# Improving the detection and management of non-communicable diseases among the adult population of the catchment areas of a rural primary healthcare unit in Sidama National Regional State, Ethiopia: A study protocol

**DOI:** 10.1101/2024.04.11.24305685

**Authors:** Melaku Haile Likka, Hiwot Abera Areru, Betelihem Eshetu Birhanu, Desalegn Tsegaw Hibistu, Bernt Lindtjørn

## Abstract

**Background:** The World Health Organization has designed a package of essential non-communicable diseases (PEN) strategy to improve the detection and management of NCDs. However, the implementation of the PEN in Ethiopia is at an early stage and the readiness of rural primary healthcare units (PHCUs) to implement the strategy is unknown. We, therefore, propose to apply the strategy in the catchment areas of Dobe-Toga Health Center, a rural PHCU in Sidama National Regional State (SNRS), Ethiopia, and improve the NCDs care among adults aged≥45 years.

**Aim:** We aim to determine the prevalence of undiagnosed hypertension, pre-T2DM mellitus, T2DM, and comorbidity of hypertension and T2DM among the older adults in the study areas, apply the WHO-PEN-based care model for the participants diagnosed with T2DM and/or hypertension and evaluate its effectiveness in controlling blood glucose and/or pressure. The readiness of PHCUs to implement the WHO-PEN approach in the region will also be determined. Additionally, we examine the influence of diagnosis with hypertension and/or T2DM on the willingness of the study participants to join and/or renew membership in community-based health insurance (CBHI).

**Methods:** The study will be conducted in catchment areas of Dobe-Toga Health Center from April to November 2024. A cross-sectional survey of 41 health centers and 4 primary hospitals, triangulated with qualitative data, will be employed to assess the readiness of the PHCUs to implement the WHO-PEN interventions while the qualitative data for this study has not been collected yet, the quantitative data was collected through observation checklist of inputs. The remaining studies will be conducted in two phases. In Phase 1, cross-sectional surveys will be conducted to determine the prevalence of undiagnosed hypertension, T2DM, pre- T2DM, and comorbidity of T2DM and hypertension in a randomly selected sample of 3301 older adults. Additionally, the participants’ willingness to pay (WTP) for HbA1c tests will be assessed, and CBHI-related surveys will be conducted. In the second phase, the cohorts will be linked to the health center and will receive the WHO-PEN-based care model. In phase 2, the effects of the care model in controlling blood pressure and glucose will be examined. Furthermore, the adherence to self-care practices of the cohorts will be determined.

## 1. Introduction

### 1.1. Background

Chronic (non-communicable) diseases, including cardiovascular diseases (such as heart attacks, hypertension, and stroke), cancers, chronic respiratory diseases (such as chronic obstructive pulmonary disease and asthma), and diabetes, are conditions characterized by long duration and caused by a combination of genetic, physiological, environmental, and behavioral factors. Non-communicable diseases (NCDs) are the leading causes of premature death and disability globally [1,2].

Hypertension and diabetes mellitus are the two common NCDs, particularly in low-and-middle-income countries (LMICs) [3]. Globally, around 1.28 billion adults aged 30 to 79 have hypertension, with two-thirds residing in LMICs. The number of adults with hypertension increased from 594 million in 1975 to 1.13 billion in 2015. The African Region has the highest prevalence of hypertension at 27% [4]. The other major NCD, diabetes mellitus (DM), affects more than half a billion adults worldwide between the ages of 20 and 79, accounting for 10.5% of all adults in this age range. By 2030, it is predicted that the number of adults with diabetes, predominantly type 2 diabetes mellitus (T2DM), will reach 643 million, and by 2045, it will increase to 783 million [5]. In Sub-Saharan Africa (SSA), the prevalence of overall diabetes and pre- diabetes were 6.8% and 25%, respectively. The adult population of 50-59 years old is more affected by DM, and the prevalence has been reported to be 14.9% [6]. The World Health Organization (WHO) reported that diabetes was ranked as the seventh cause of death in 2016, accounting for 1.6 million deaths in that year, where half of them were people under the age of 70 years [7,8].

Ethiopia faces a triple health challenge: NCDs, injuries, and communicable diseases [9]. The 2018 Ethiopian NCD and Injury Commission Report pointed out that NCDs share 37.5% of the disease burden [10]. Nearly 40% of deaths in Ethiopia are due to NCDs [11], with hypertension and diabetes being responsible for over a third of the disease burden and 43% of fatalities [12,13]. Hypertension is a significant public health concern in Ethiopia, responsible for over half of cardiovascular diseases and affecting nearly a quarter of the population [12]. Controlling hypertension lowers the risk of developing other NCDs, such as heart failure, chronic kidney disease, and stroke [14]. However, only 20% of the adults were able to control their hypertension in Ethiopia [4]. In addition, the pooled prevalence of uncontrolled hypertension was 48% in Ethiopia [15]. This highlights a significant gap in the diagnosis and treatment of hypertension in Ethiopia.

Similarly, the prevalence of DM was 6.5% in Ethiopia [16], and that of undiagnosed diabetes ranges from 5.80% to 10.2% [17,18]. These studies determined the prevalence of undiagnosed diabetes using less reliable techniques such as fasting and random blood sugar tests. None of the studies on the prevalence of DM used hemoglobin A1c (also called HbA1c, glycated hemoglobin test, or glycohemoglobin). This blood test provides an average over the past 2 to 3 months of blood sugar (glucose) for the diagnosis of diabetes mellitus. Glycated hemoglobin HbA1c is considered the gold standard for assessing the compensation and treatment of diabetes and diagnosis of DM [19]. Furthermore, for the early diagnosis of T2DM and pre-diabetes in asymptomatic persons, the American Diabetes Association and the WHO recommend using HbA1c with a cut-off value of 6.5% [20].

The WHO has designed evidence-based recommendations, the package of essential non-communicable diseases (PEN), to improve the detection, treatment, and management of NCDs such as diabetes and hypertension. These packages include affordable drugs for treatment, non-pharmacological and pharmaceutical techniques for modifying risk factors, and cost-effective strategies for early NCD detection [2]. Early detection and management are critical interventions for preventing hypertension and diabetes complications and deaths, as well as improving treatment outcomes. However, nearly one in every two people has undiagnosed diabetes, the majority of whom are type 2 patients around the globe [21].

Ethiopia has taken measures to address the problem of major NCDs such as hypertension and diabetes. It has developed a national guideline that promotes the early detection and treatment of NCDs and has worked to enhance the capacity of primary healthcare units (PHCUs) in low-resource settings to integrate and scale up the treatment of these diseases [2,22–24]. The PHC is an essential tool for addressing the rising burden of NCDs and is the best method for providing integrated and equitable NCD care [25].

However, the utilization and quality of service remain inadequate in Ethiopia, owing to insufficient technical capacity for NCD detection, care, and management, a lack of diagnostic tests, little support from national authorities, limited equipment and medicines, and inadequate integration of NCD care to the PHCUs [22,26,26]. Furthermore, the implementation of the PEN in Ethiopia is at an early stage, and the readiness of the PHCUs to implement the WHO-PEN disease intervention strategy is unknown.

We, therefore, propose to improve the early detection and management of hypertension, pre-T2DM, and T2DM among older adults (aged 45 years and older) in the catchment area of Dobe-Toga Health Center, a rural PHCU in Sidama National Regional State (SNRS), Ethiopia. These will be achieved by undertaking community-based screening and implementing a care model detailed in Section 2.5 for the cohorts of individuals diagnosed with the above conditions. As part of implementing the care model, we will determine the prevalence of undiagnosed hypertension, type-2 diabetes mellitus (T2DM), pre-T2DM, and the burden of comorbidity of hypertension and T2DM among the study participants. We will also examine the effectiveness of the care model in reducing the level of blood pressure and hemoglobin A1c after implementation of the care mode for six months. The readiness of the PHC system in SNRS to implement the WHO-PEN approaches will be determined in this study.

Furthermore, we will determine whether having a family member diagnosed with one or more of the abovementioned conditions will affect the respondents’ decision to renew their membership and enroll in the community-based health insurance (CBHI) scheme implemented in Ethiopia in 2014 [27]. Also, we will determine the willingness of the participants to raise the annual premium for membership in case the schemes face sustainability challenges due to moral hazards and adverse selections. Unlike most schemes in LMICs [28], the Ethiopian CBHI does not apply co-payment for service delivery to eligible individuals, which is crucial in reducing the impact of moral hazards. We will examine the willingness of the CBHI-member respondents to co-pay for healthcare services.

### 1.2. Study aims

The study aims to apply the WHO-PEN-based care model to the elderly population (aged 45 years and above) living in the catchment areas of Dobe-Toga Health Center, a rural PHCU in Shebedino District of SNRS, and determine the prevalence of undiagnosed hypertension, pre-T2DM, T2DM and comorbidity of both hypertension and T2DM in this population. We will also evaluate the effectiveness of the care model in controlling blood pressure and glucose levels. Additionally, we will assess the rural PHCUs’ readiness to implement the WHO PEN disease intervention approach in the region and examine the influence of hypertension and T2DM on the willingness to join and renew membership of CBHI among the study participants.

### 1.3. Objectives of the study

The study is divided into 3 categories, each with its objective(s):

#### a) Studies on hypertension

1. To determine the prevalence of undiagnosed hypertension among the adult population in the catchment kebeles of Dobe-Toga Health Center, Shebedino District, SNRS.
2. To examine the effect of implementing WHO-PEN-based hypertension and T2DM management and care package in reducing the level of blood pressure among the study participants in the study area.
3. To determine the overall level of self-care practice among hypertensive clients on follow-up and factors that determine the level of self-care practice among the study participants.

#### b) Studies on T2DM

4. To determine the prevalence of undiagnosed T2DM using HbA1c as a confirmatory diagnosis among the adult population in the study areas.
5. To assess the effect of the WHO-PEN approach on the change in blood sugar level, as measured by HbA1c, among adults diagnosed with T2DM over six months in the study area.
6. To determine adherence to self-care practices among adults diagnosed with T2DM in the study areas.
7. To determine the magnitude of willingness to pay for HbA1c testing for the diagnosis of blood sugar levels in the catchment areas of Dobe-Toga Health Center, Shebedino District, SNRS.

#### c) Other studies

8. To determine the readiness of PHCUs in SNRS to implement the WHO-PEN intervention.
9. To determine the prevalence of undiagnosed pre-T2DM among the adult population in the catchment areas of Dobe-Toga Health Center, Shebedino District, SNRS.
10. To determine the burden of comorbidity of hypertension and T2DM in the study area.
11. To compare the families with members with and without NCDs to enroll in CBHI in the study areas.
12. To compare the families with members with and without NCDs to renew the CBHI membership in the study areas.
13. To determine the willingness for co-payment for health services among the study participants in the study area
14. To assess the willingness to raise the annual membership premiums as an option to sustain the scheme in the community, 2024

## 2. Methods and designs

### 2.1. Study setting and period

The study will be conducted in catchment areas of a rural PHCU called Dobe-Toga Health Center, Shebedino District, one of the 36 districts in SNRS, from April to November 2024. The district’s capital is Leku town, located about 300 kilometers south of Addis Ababa, the nation’s capital, and 24 km from Hawassa, the capital city of SNRS. According to the Ethiopian Central Statistical Agency’s population projections for 2021/2022, the district’s total population was 204,618 (male 100,263 and female 104,355). The district has 23 rural and nine urban kebeles (the smallest administrative unit in Ethiopia) with six health centers, including Dobe-Toga Health Center. Each rural health center has 3-4 satellite health posts collaborating with a primary hospital to form the PHCUs. Dobe-Toga Health Center is serving 38,874 people in its catchment area of four kebeles. According to the age distribution of SNRS, adults aged ≥45 constitute 13%, equaling 5054 individuals.

### 2.2. Study designs

A cross-sectional survey of samples of PHCUs (health centers and primary hospitals), triangulated with qualitative data, will be employed to assess the readiness of the PHCUs in SNRS to implement the WHO-PEN interventions (Objective 8). The qualitative data to triangulate the cross-sectional survey will be obtained through in-depth interviews of NCD focal persons of selected PHCUs and healthcare authorities in the SNRS Health Bureau and selected lower-level administrative organs.

The remaining studies will be carried out in two phases. In the first phase, community-based cross-sectional surveys will be conducted to identify undiagnosed hypertension and T2DM cohorts. The cohorts will be linked to Dobe-Toga Health Center for follow-up and receive the WHO-PEN- -based Hypertension and Diabetes Mellitus Care Model described in the <u>subsequent section</u>. The <u>Care Model</u> will be intervened for 6-8 months. In addition to identifying the cohorts, CBHI-related surveys will be conducted on the same study population during the community-based survey and at the end of the follow-up. The follow-up tests and examinations will be performed during the 1^st^, 3^rd,^ and 6^th^ months of diagnosis at the health center. A pre-post design will be employed for the second phase of the studies.

### 2.3. Population

For the community-based cross-sectional studies, all adults aged ≥45 living in the catchment areas of Dobe-Toga Health Center and their households will be considered source populations. For the cohort study: adults whose age ≥45 years, living in the catchment areas of Dobe-Toga Health Center who will participate in the community-based cross-sectional study, and diagnosed with raised blood pressure and diabetes mellitus using a fasting blood sugar (FBS) test will be the source population. For the cross-sectional surveys of PHCUs, all health centers and primary hospitals in SRS will be considered the source population.

### 2.4. Eligibility Criteria

For the community-based cross-sectional studies, adults (age 45 or above) who have lived in the selected kebeles for at least six months will be eligible. For the cohort studies, individuals diagnosed with hypertension and T2DM (first screened with FBS, then confirmed with HbA1c test) will be eligible. Health centers and primary hospitals in SNRS will qualify for the surveys of PHCUs. Individuals with severe cognitive challenges and disability and pregnant women will be excluded from the study as they may temporarily have hypertension and diabetes due to their physiological processes.

### 2.5. The Hypertension and Diabetes Mellitus Care Model Package

The intervention for the pre-pos study at Dobe-Toga Health Center is named “Hypertension and T2DM- Care Model”. This care model package includes community-based screening for hypertension, pre-T2DM, and T2DM; strengthening the capacity of the health center by training the staff; proper diagnosis of Hypertension, pre-T2DM, and T2DM; improving the patient ownership; and follow-up system through clinical and laboratory-based management, drug treatment, and behavioral counseling at the point of care at the Dobe-Toga Health Center. The care model aims to improve the detection and management of hypertension, T2DM, quality of life, adherence to the care model, patient ownership, and diabetes mellitus control.

#### Community-Based Screening for hypertension, pre-T2DM and T2DM

To carry out the community-based screening to map the burdens of undiagnosed hypertension, pre-T2DM, and T2DM among the adult population, we will carry out the following activities: (1) identifying the target population and location for the screening program; (2) developing a screening protocol that includes measurement of blood glucose levels and other relevant clinical and demographic information; (3) recruiting and training the local PHCU health professionals/data collectors to conduct the screening, including providing them with the necessary equipment and supplies (4) carrying out the screening program in the community, reaching as many eligible individuals as possible; (5) collecting and analyzing the screening data to identify the prevalence of undiagnosed hypertension, pre-T2DM, and T2DM in the community as well as any risk factors, and linking the diabetic clients to Dobe-Toga Health Center for further management and care.

#### Strengthening the capacity of the health center to detect hypertension, pre-T2DM and T2DM by training the staff

The following activities will be carried out or will be developed to strengthen the capacity of the target PHCU (1) Designing and up-to-date training document that covers the basics of hypertension and diabetes detection and management that takes into account the current guidelines, such as the WHO-PEN intervention approach [2], best practices, and evidence-based recommendations about the detection and management of the conditions; (3) Identifying and selecting trainers with expertise in diabetes and hypertension care to serve as trainers, and recruiting trainees; (4) Conducting the training sessions using a combination of classroom instruction, hands-on training, and mentoring; (5) Providing ongoing support such as refreshment training to primary health professionals to ensure that they can effectively detect and manage diabetes in their patients; (6) Monitoring and evaluating the progress of the primary healthcare professionals in detecting and managing hypertension, T2DM, and pre-T2DM.

#### Proper diagnosis of hypertension, pre-T2DM, and T2DM

Concerning the adequate diagnosis of hypertension pre-T2DM, and T2DM as part of the care model, the following procedures will be taken into consideration: (1) Obtain a detailed patient history: Inquire about symptoms and risk factors for hypertension pre-T2DM, and T2DM, such as family history, sedentary lifestyle, overweight/obesity, age, etc.; (2) provide blood pressure measurement and blood glucose testing to determine if the patient is hypertensive and hyperglycemic, hypoglycemic, pre-hyperglycemic or normal. A fasting blood sugar (FBS) test will be used to screen possible pre-T2DM and T2DM cases; (3) confirmation of the possible pre-T2DM and T2DM cases using HbA1c test [29]; (4) undergoing additional biochemical tests, including urine tests, lipid tests, kidney function tests, foot exams, etc. to aid in the treatment, and monitor complications.

#### Improving patient ownership

Patient ownership or patient engagement or self-management as part of the care package is essential in managing hypertension, pre-T2DM, and T2DM. Some of the strategies that will be included under the care package to improve patient ownership will be: (1) Providing patients with accurate and comprehensive information about hypertension, pre-T2DM, and/or T2DM and their management can help them understand the condition and take an active role in managing it (education); (2) Encouraging patients to set specific, measurable, and achievable goals related to their diabetes management can help them stay motivated and focused (goal setting); (3) Focusing on the patient’s individual needs, preferences, and values can help build trust and improve communication between the patient and healthcare provider (patient-centered care); (4) Involving patients in decisions about their care can help them feel more invested in the process and more likely to adhere to treatment plans (collaborative decision-making); (5) Encouraging patients to regularly monitor their blood sugar levels and other diabetes-related parameters can help them identify patterns and make adjustments to their treatment plans as needed (self-monitoring); (6) Connecting patients with others who have T2DM can provide a sense of community and support, which can be beneficial for improving self-management (support groups); (7) Setting up phone call reminders for patients to take their medications can help to ensure that they remain adherent to their prescribed regimen and it facilitates self-management, communication, and education [30].

### 2.6. Sample Size Determination

For the community-based cross-sectional surveys, the sample size has been calculated using OpenEpi version 3.01 [31], and the following parameters: p=33.3% (the weighted prevalence of undiagnosed hypertension in Wolaita Zone, Ethiopia was 29.8%% (26.5%-33.3%) [32]. To get a larger sample size, we have taken the upper limit of this prevalence (33.3%)), design effect (DEFF) = 2, confidence level (CL)=95%, non-response rate (NRR)=10%, and adjusted for finite pop correction (N=5054). Accordingly, the sample size has been calculated to be 3301. All the community-based survey (prevalence of hypertension, pre-T2DM, T2DM, comorbidity of both T2DM and hypertension and CBHI) data will be collected from these individuals.

For the pre-post-study design to be implemented at the health center (second phase), all the individuals diagnosed with hypertension and T2DM will be followed up and included in the studies.

For the quantitative data for the study to assess the readiness of the PHCUs in SNRS to implement WHO-PEN, we sampled 30% of the PHCUs in the region. SNRS has a total of 150 PHCUs—137 health centers and 13 primary hospitals. Among them, 41 health centers and four primary hospitals (45 PHCUs) were selected randomly and included in the study. The qualitative data to triangulate the readiness findings will be obtained via in-depth interviews with regional and lower-level administrative healthcare managers and the NCD focal persons at the selected district health offices.

### 2.7. Sampling Procedure and Follow-up of the Schedule

A multi-stage sampling technique will be applied to select the study participants. First, one rural PHCU— Dobe-Toga Health Center with its four catchment kebeles—has been selected considering its total population size and the fact that all the catchment kebeles are located rurally among the six health centers and 23 kebeles in the district. The health center’s catchment kebeles are Gonowa-Gabalo, Dobe- Toga, Howolso, and Gobe-Hebisha, for a total of 38,874 population living in 7,933 households. Then, the sample will be allocated to the four kebeles in proportion to the size of the study population. The households with the relevant study participants in each kebele will be selected through the systematic sampling technique using the list of households as a sampling frame. The K value for systematic sampling will be determined by dividing the total adult population by the estimated sample size, and then every K household will be selected (Figure 1). However, if there are more than eligible respondents, all eligible individuals within the selected household will be considered. As a result, the final sample size may exceed the calculated one.

**Figure 1:**
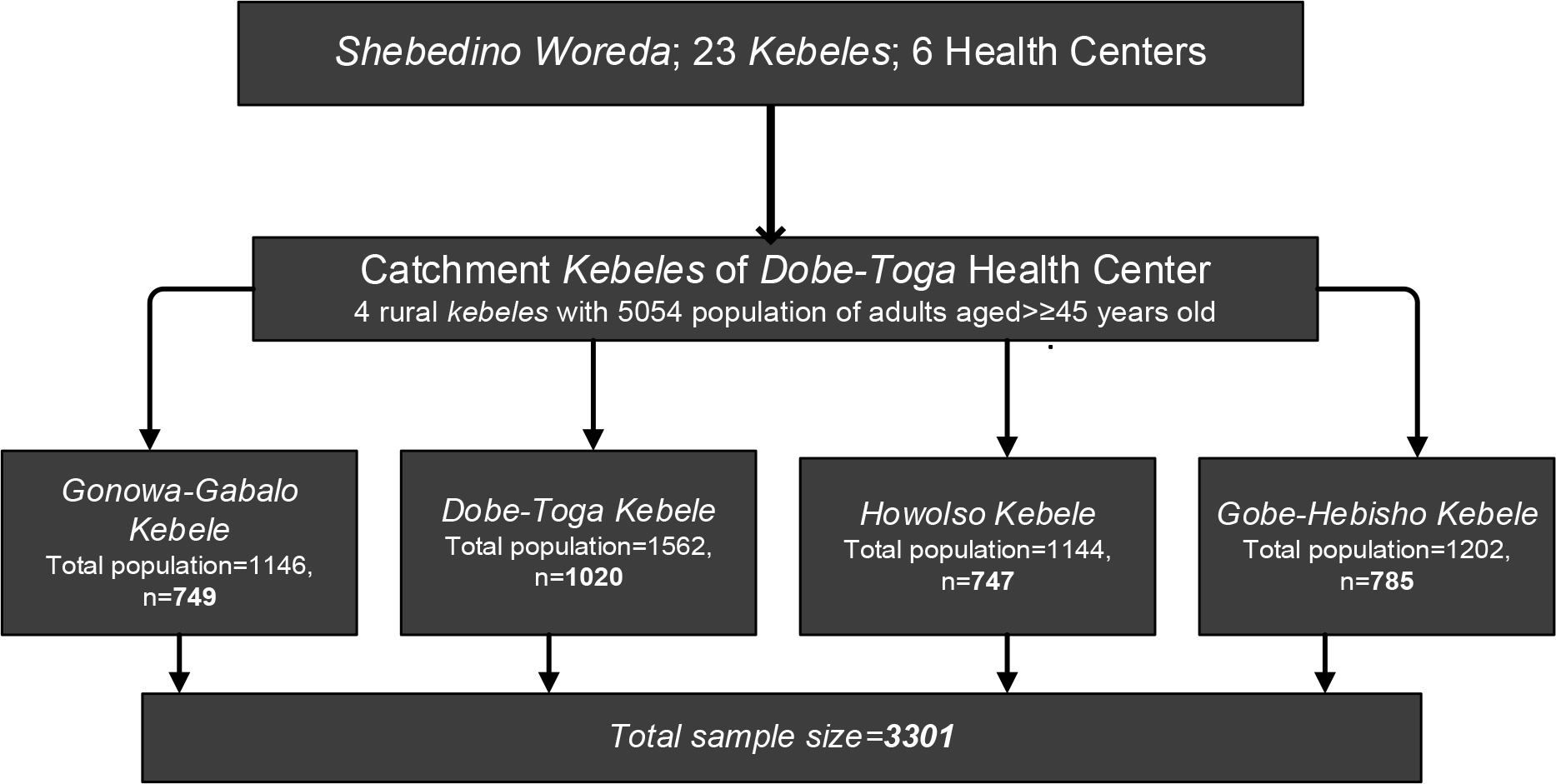
Schematic presentation of the sampling procedure.

Following the selection of the kebeles, a cross-sectional study will be conducted to determine the burden of hypertension, T2DM, pre-T2DM, and comorbidity of both T2DM and hypertension among the participants. While those diagnosed with hypertension will be declared positive without further assessment, those who will be diagnosed to have pre-T2DM and T2DM will be ascertained using an HbA1c test. Finally, the individuals diagnosed with hypertension and T2DM (ascertained with HbA1c) will be linked to Dobe-Toga Health Center for further care and application of the Care Model specified earlier for 6-8 months. In addition, the individuals will also be asked about their willingness to pay (WTP) for HbA1c tests for detection and hypertension. The participants will also be asked about their participation in CBHI and willingness to pay (WTP) for copayment and raise the annual premiums to sustain the CBHI program in their community. To compare the decisions about the CBHI, the participants will also be asked about the CBHI-related items at the end of the follow-up.

### 2.8. Study variables

#### The outcome variables

NCDs care and management input and process indicators of health centers and primary hospitals, systolic and diastolic blood pressure and blood glucose level during the community-based screening, 3rd and 6th months of intervention and follow-up; HbA1c during community-based screening (for those whose FBS≥5.6 millimole/liter mmol/L) of blood only), 3rd and 6^th^-month follow-up; self-care practice; adherence to the WHO-PEN-based care model; WTP for HbA1c for diagnosis of T2DM during the community-based screening; enrolment status of the community-based health insurance (during the community-based screening and at the end of the follow-up); willingness to renew CBHI membership; willingness to raise the annual premium for CBHI membership; willingness to pay the co-payment during healthcare services.

#### The exposure variables

Locations of the facilities to be assessed for the readiness of the PHCUs; type of the facilities; managing authorities of the health facilities; area of the facility (urban/rural); types of services provided; socio-demographic and economic characteristics (sex, age, marital status, educational level, occupation, income/wealth status); self-rated health status; family size; wealth index of the households; distance to the health facility, behavioral risk factors (tobacco and khat use, alcohol consumption); dietary behaviors (fruit and vegetable intake, salt intake, and physical activity); history of NCDs; physical measurements (height, weight, body mass index, cholesterol level, and blood pressure).

### 2.9. Measurements of the variables

The WHO’s Service Availability and Readiness Assessment (SARA) [33] methodology and WHO-PEN Interventions for Primary Healthcare reference manuals will be used to assess the readiness of the PHCUs to implement the WHO-PEN intervention.

Hypertension is the average blood pressure (BP) of three measures is >140/90 mmHg or >130/80 mmHg for patients with diabetes or chronic kidney disease [34]. The professional data collector will make sure the following before measuring the BP:

- the patient rests for at least 5 minutes before the measurement and avoided alcohol, caffeine, and physical activity;
- a properly calibrated automatic BP monitor (Omron HEM 7080 BP apparatus) is being used;
- the patient sits with their back supported and their arm supported at heart level;
- three consecutive BP readings will be taken at least five minutes apart with an appropriate-sized cuff
- Blood pressure readings will be recorded and averaged to measure an individual’s blood pressure status statistically.

The level of self-care practice among hypertensive clients on follow-up will be assessed using the Hypertension Self-Care Activity Level Effects (H-SCALE). This 31-item scale measures self-care aspects, including medication adherence, dietary management, smoking status, physical activity, weight management, and alcohol intake [35,36]. The level will be categorized as good self-care practice (adherence to all the components of the H-SCALE) and poor self-care practice (non-adherence to at least one element of the H-SCALE).

Type-2 diabetes mellitus will be confirmed if one of the following criteria is fulfilled: a positive history of diabetes mellitus, the use of anti-diabetic medication, and fasting blood sugar (FBS) at the community-based screening. HbA1c will confirm the result at the rural PHC facility. The HbA1c test evaluates the average blood sugar (glucose) levels in the red blood cells over the previous two to three months. It is used to diagnose and monitor diabetes and determine the risk of acquiring diabetes and other long-term problems, making it a crucial component of diabetes care [37,38]. According to the ADA, pre-diabetes (Pre-DM) will be considered if an FBS between 5.6 mmol/L to 6.9 mmol and HbA1c 39–47mmol/mol while the T2DM will be confirmed if the FBS≥7.0 mmol/L and the HbA1c ≥48 mmol/mol. The HbAlc is preferred to other diagnosis methods due to its superior advantages, like greater convenience (fasting not required), greater pre-analytical stability, and fewer day-to-day perturbations during stress, changes in diet, or illness [29,39].

To assess willingness to enroll in CBHI and renew their membership of the schemes, between families with members with and without NCDs, CBHI membership and renewal-related items will be administered to all of the study participants during the initial community-based survey and at the end of the follow-up time. The initial assessment will determine the differences in willingness to participate in the CBHIs among participants with and without hypertension, T2DM, pre-T2DM, and disability. The second phase of the assessment will evaluate the differences in the willingness to participate or renew membership before and after diagnosis with the conditions. In both cases, those who reply “no” will be classified as non-CBHI members/not willing to renew the membership. Those who say “yes” will be considered CBHI members willing to renew their membership. In all cases, explanatory questions such as reasons for their preferences in membership, their assessment of the quality of care in the catchment areas, etc., will be assessed.

To assess the WTP extra premiums for annual CBHI membership and copayment for medical services, we will employ the double-bound dichotomous choice-contingent method (DBDC-CVM), one of the techniques direct methods used to illicit individuals’ WTP for public and non-market goods or services [40]. This elicitation method will be used since it is information-intensive, asymptotically more efficient, less sensitive to the starting point and strategic biases, and reduces the demand for a large sample size [40,41]. Suppose that the indirect utility of an individual depends on buying a health insurance policy and income y. Let q_1_ and q_0_ measure the level of utility associated with and without health insurance, respectively; WTP is the amount of money an individual is WTP as a premium, X represents the vector of other factors (such as age, sex, education, health status, etc.) that may affect the preferences of individuals, π shows the perceived probability of falling sick and Ɛ captures other factors that are unobservable to the researcher. Then, the individuals will buy the health insurance policy only if v [(q_1_, y- WTP, X, π) + Ɛ_1_) ≥ v [(q_0_, y, X, π) + Ɛ_0_); where: Ɛ_1_ and Ɛ_0_ are random errors distributed independently with mean zero.

In DBDC-CVM, each respondent is asked if s/he is willing to pay the first bid. If s/he says ‘yes’ to the first bid, a second higher bid is given, and their WTP is asked. A second lower bid is provided if s/he says ‘no’ to the initial bid. If s/he says ‘no’ or ‘yes’ to both the first and the second bids, then s/he is asked to mention the maximum amount of money that s/he is willing to pay, as illustrated by Figure 2 [40].

**Figure 2:**
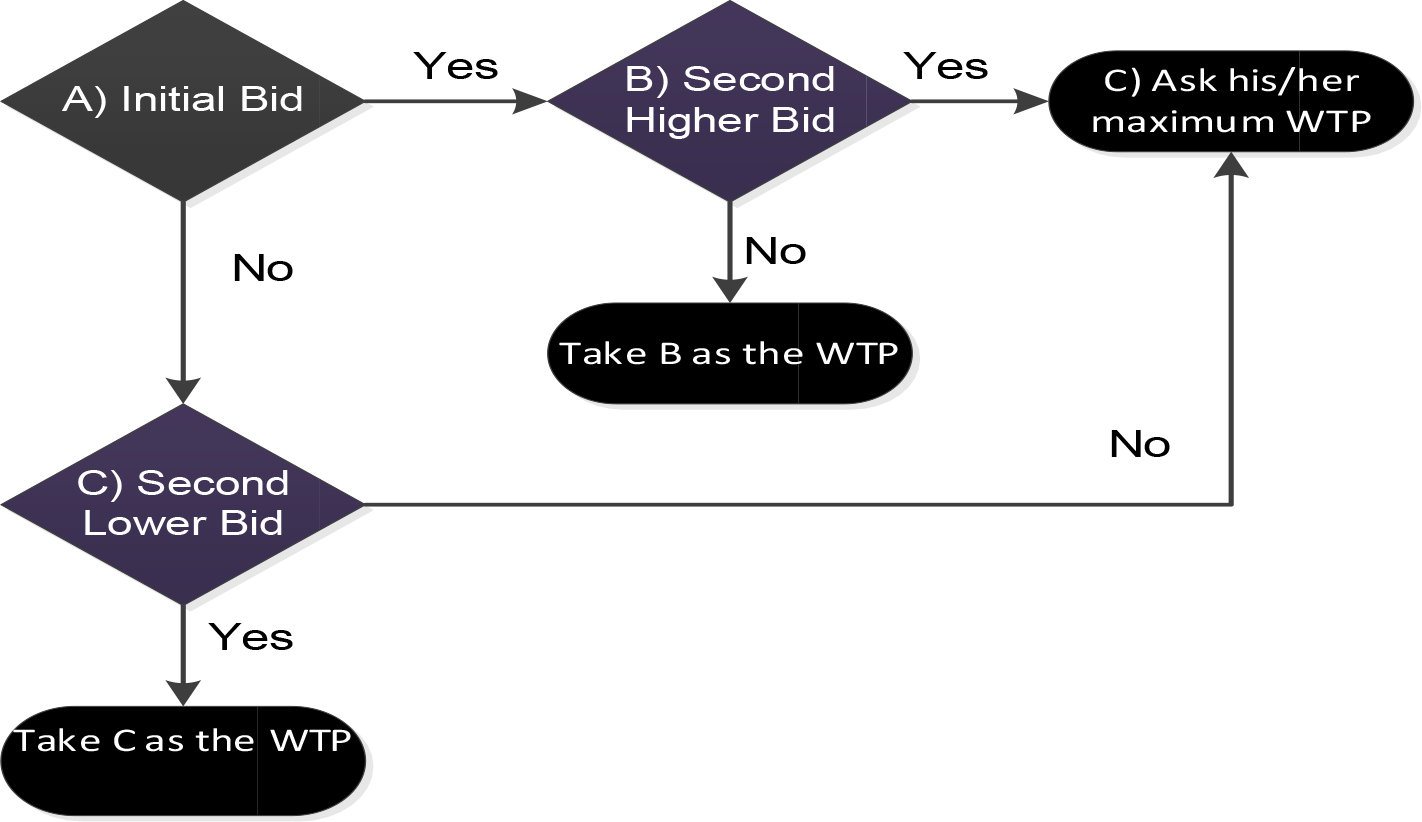
The double-bound dichotomous choice-contingent valuation method will be used to elicit WTP for extra premium and copayment in the study area.

Accordingly, for this study, four initial bids for extra annual premiums and co-payments were set for the respondents to choose from using the lottery method during the survey. The second higher and lower bids will be twice and half of the initial bids chosen. These initial bids will be 200, 250, 300, and 400 Ethiopian birrs (for the annual premiums) and 10%, 15%, 20%, and 25% for the copayments. Before eliciting the WTP, hypothetical situations will be presented. As the purpose of the WTP study is to sustain the CBHI, we will ask about their preference between the discontinuation of the schemes and the increase in the annual premiums and copayments for the services presented above.

A DBDC-CVM will be used to assess the WTP for HbA1c diagnosis of T2DM (Figure 2). Four initial bids (250, 300, 400, and 500 Ethiopian birrs) will be randomly assigned to participants. The second highest and lowest bids will be 150% and 50% of the initial bids, respectively.

The measurement of the exposure variables will be detailed during the publication of the research report.

### 2.10. Statistical Analysis

After the data are collected using the KoboToolbox, they will be exported to R version 4.3.3 and STATA version 16 for relevant statistical analyses. Before analyzing the data, data cleaning will be undertaken, including removing errors, inconsistencies, or missing data from the dataset. Then, the main characteristics of the data will be summarized. Binary logistic regression will be utilized for the community–based study to identify factors associated with the prevalence of undiagnosed hypertension, T2DM, pre-diabetes, and double burden of hypertension and T2DM.

The change in mean blood pressure, HbA1c, and blood glucose level at 3 and 6 months will be determined using a dependent t-test to measure the effect of the exposure variables on the outcome variables. Linear regression will be carried out to determine the predictors for improving blood pressure and blood glucose levels and for the change in HbA1c. The exposure variables will be checked for the multi-collinearity effect using variance inflation (VIF). The strength of the association will be presented with a 95% confidence interval (CI) with the corresponding adjusted odd ratio (AOR). A p-value of < 0.05 will be taken as a statistical significance. To determine adherence to the anti-hypertensive and anti- diabetic medications and care, ordinal logistic regression will be employed.

Binary logistic regression will be used to assess the relationship between membership to CBHI and willingness to renew the membership. To determine the WTP for HbA1c for T2DM diagnosis, additional premium and copayment using DBDC-CVM, seemingly unrelated bivariate probit regression will be used, and to determine the mean/median WTP, a STATA module called WTPCIKR64 will be used to determine Krinsky and Robb’s Confidence Interval for Mean/median WTP. The multivariate analysis plans for each objective of the study have been summarized in Table 1

**Table 1:**
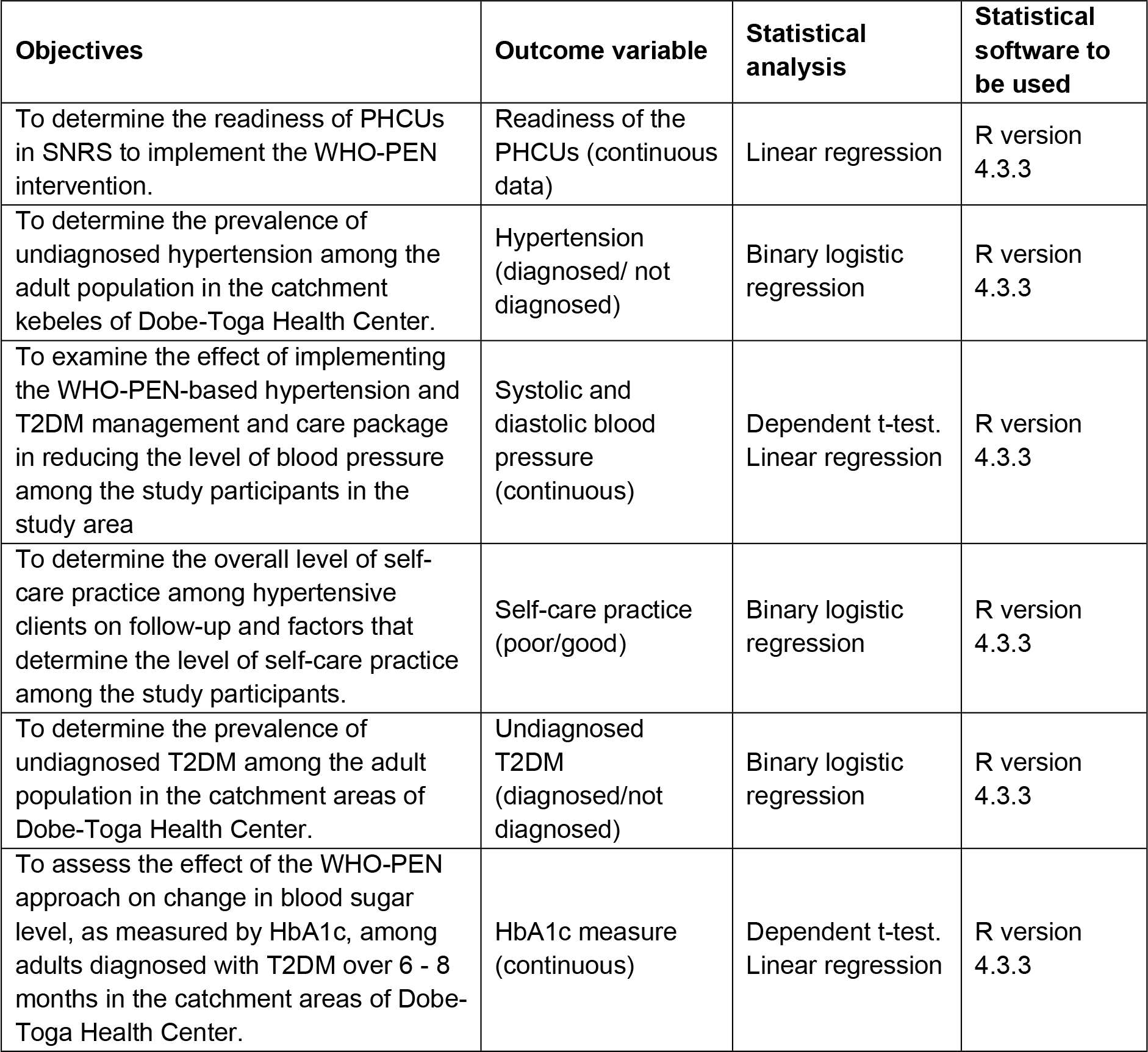
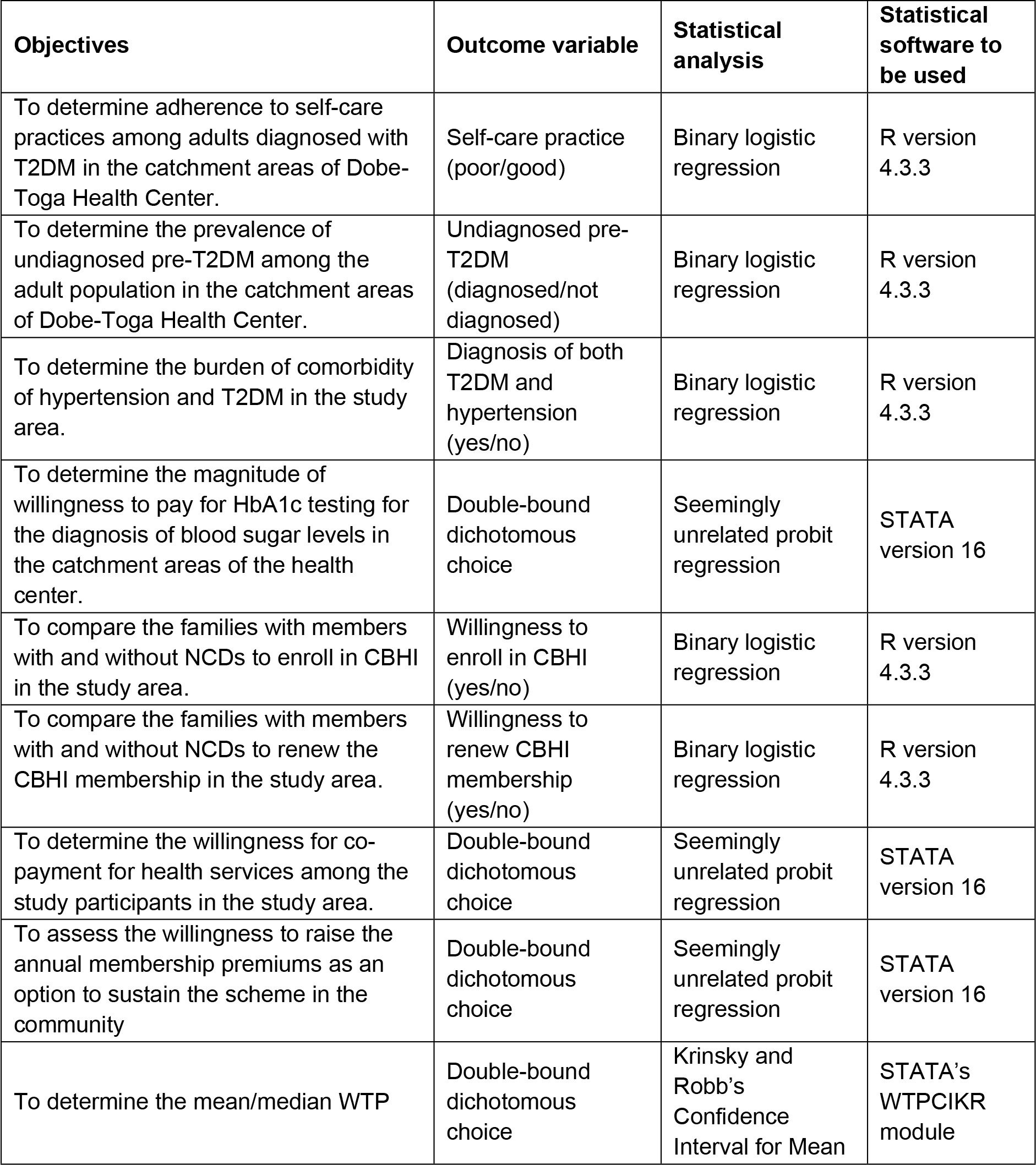
Statistical analysis plans for each objective in this research.

| **Objectives** | **Outcome variable** | **Statistical analysis** | **Statistical software to be used** |
| --- | --- | --- | --- |
| To determine the readiness of PHCUs in SNRS to implement the WHO-PEN intervention. | Readiness of the PHCUs (continuous data) | Linear regression | R version 4.3.3 |
| To determine the prevalence of undiagnosed hypertension among the adult population in the catchment kebeles of Dobe-Toga Health Center. | Hypertension (diagnosed/ not diagnosed) | Binary logistic regression | R version 4.3.3 |
| To examine the effect of implementing the WHO-PEN-based hypertension and T2DM management and care package in reducing the level of blood pressure among the study participants in the study area | Systolic and diastolic blood pressure (continuous) | Dependent t-test.<br>Linear regression | R version 4.3.3 |
| To determine the overall level of self-care practice among hypertensive clients on follow-up and factors that determine the level of self-care practice among the study participants. | Self-care practice (poor/good) | Binary logistic regression | R version 4.3.3 |
| To determine the prevalence of undiagnosed T2DM among the adult population in the catchment areas of Dobe-Toga Health Center. | Undiagnosed T2DM (diagnosed/not diagnosed) | Binary logistic regression | R version 4.3.3 |
| To assess the effect of the WHO-PEN approach on change in blood sugar level, as measured by HbA1c, among adults diagnosed with T2DM over 6 - 8 months in the catchment areas of Dobe-Toga Health Center. | HbA1c measure (continuous) | Dependent t-test.<br>Linear regression | R version 4.3.3 |
| To determine adherence to self-care practices among adults diagnosed with T2DM in the catchment areas of Dobe-Toga Health Center. | Self-care practice (poor/good) | Binary logistic regression | R version 4.3.3 |
| To determine the prevalence of undiagnosed pre-T2DM among the adult population in the catchment areas of Dobe-Toga Health Center. | Undiagnosed pre-T2DM (diagnosed/not diagnosed) | Binary logistic regression | R version 4.3.3 |
| To determine the burden of comorbidity of hypertension and T2DM in the study area. | Diagnosis of both T2DM and hypertension (yes/no) | Binary logistic regression | R version 4.3.3 |
| To determine the magnitude of willingness to pay for HbA1c testing for the diagnosis of blood sugar levels in the catchment areas of the health center. | Double-bound dichotomous choice | Seemingly unrelated probit regression | STATA version 16 |
| To compare the families with members with and without NCDs to enroll in CBHI in the study area. | Willingness to enroll in CBHI (yes/no) | Binary logistic regression | R version 4.3.3 |
| To compare the families with members with and without NCDs to renew the CBHI membership in the study area. | Willingness to renew CBHI membership (yes/no) | Binary logistic regression | R version 4.3.3 |
| To determine the willingness for co-payment for health services among the study participants in the study area. | Double-bound dichotomous choice | Seemingly unrelated probit regression | STATA version 16 |
| To assess the willingness to raise the annual membership premiums as an option to sustain the scheme in the community | Double-bound dichotomous choice | Seemingly unrelated probit regression | STATA version 16 |
| To determine the mean/median WTP | Double-bound dichotomous choice | Krinsky and Robb's Confidence Interval for Mean | STATA's WTPCIKR module |

### 2.11. Quality Assurance

Appropriate actions are being taken to maintain the research’s quality, from designing the tool to data analysis and interpretations. The questionnaires have been adapted from standard data collection instruments used by the WHO STEP-wise approach surveillance for non-communicable diseases in English and translated into the local language, Sidaamu Affoo. The translated Sidaamu Affoo version will be translated back to the original language by other bilingual experts to check the consistency of the tool. Data will be collected through face-to-face interviews using an interviewer-administered Sidaamu Affoo version of the questionnaire using the Android-based KoboToolbox platform installed on smartphones [42].

The tool was pretested in Remeda Kebele of Habela District, SNRP, with 160 individuals (5% of the sample size). Appropriate modifications have been made following the findings of the pre-test. Data collectors and supervisors were recruited by considering their level of education, type of profession, experience in data collection, and language ability. Before data collection was commenced, data collectors and supervisors were trained on utilizing the KoboToolbox data collection platform, experience exchange, and demonstrations on using the KoboToolbox. The data collection procedures will be closely supervised. The Geographical Positioning System (GPS) on the KoboToolbox will be activated to see the location of each household. All questions will be checked for completeness as soon as the data collectors submit the data to the server developed for this research purpose, and immediate actions will be taken.

Proper feedback will be given to data collectors, and in some kebeles, data collection might be repeated if necessary, taking five percent of the sample size. Advanced techniques and appropriate modeling will be applied during the analysis. Important assumptions and fitness of such statistical models will be checked using standard procedures.

### 2.12. Ethical Consideration

Ethical clearance for this study was obtained in May 2023 from the Institutional Review Board (IRB) of the College of Medicine and Health Sciences, Hawassa University. Since the original protocol was modified, a revision of the ethical clearance was requested, and the Board approved the revision in April 2024.

When data collection starts, an official letter will be taken from the SNRS Health Bureau. Permission will also be requested from the Shebedino District Health Office.

The study participants will receive information about the study’s purpose and provide written consent, except for the quantitative data in Objective 8 (described in the <u>next section</u>). The informed consent will be read for those who cannot read, and if they agree to participate, a thumbprint will be taken. The study participants will also be informed about the purpose of the study. All participants’ right to self-determination will be respected. To maintain the confidentiality and privacy of the respondents, the study participants’ names will be anonymous, and the tool will be password-protected. Participants’ involvement in the study will be voluntary; participants who are unwilling to participate and those who wish to quit their participation at any stage will be told to do so without any restriction. During data collection, counseling will be provided for newly diagnosed clients.

### 2.13. Prospective recruitment of study participants

Quantitative data for Objective 8, which aims to determine the readiness of PHCUs in SNRS to implement the WHO-PEN intervention, were collected from December 1 to 12, 2023. An observation checklist of NCD inputs was used to collect the data from randomly selected 41 health centers and 4 primary hospitals in SNRS. Since the study poses minimal risks, only verbal consent was sought from the authorities of the selected health facilities. The authorities were informed about the purpose of the study and IRB-approved scripts of consent were presented to the authorities. Observations were conducted only in those facilities where verbal consent was obtained. The qualitative data for this study will be collected in April to May 2024. Informed consent will be sought from the participants of the qualitative study.

The study participants for the other objectives have not been recruited yet.

### 2.14. Dissemination Plan

The study findings will be published in international peer-reviewed Open Access journals. The potential key authors and proposed submission dates have already been identified for the study objectives. A copy of the results will be submitted and presented to the School of Public Health, College of Medicine and Health Sciences, Hawassa University; SNRS Health Bureau; and Shebedino District Health Office. The findings will also be presented to various scientific conferences and governmental and non-governmental organizations so that they can use them for planning and decision-making to improve the detection and management of NCDs.

## Author Contributions

Melaku Haile Likka, Hiwot Abera Areru, Bernt Lindtjørn, Betelihem Eshetu Birhanu, and Desalegn Tsegaw Hibistu contributed to the conceptualization and methodology. Professor Bernt Lindtjørn contributed to resource acquisition, reviewing, and editing the protocols and manuscript.

## Data Availability

No datasets were generated or analyzed during the current study. All relevant data from this study will be made available upon study completion.

## Acknowledgment

We want to acknowledge the Government of Norway for funding this research through the Norwegian Program for Capacity Development in Higher Education and Research for Development (NORHED) South Ethiopia Network of Universities in Public Health-II (SENUPH-II). We are also grateful to the Hawela District Health Office for allowing us to conduct the pre-test in one of their kebeles. We acknowledge the respondents of the pre-test in Remeda Kebele.

## Abbreviations and acronyms

ADA: American Diabetic Association
BP: Blood pressure
CVM: Contingent valuation method
DBP: Diastolic blood pressure
ETB: Ethiopian birr
FMoH-E: Federal Ministry of Health-Ethiopia
HbA1c: Haemoglobin A1c
LMICs: Low-and-middle-income countries
mmHg: Millimetre mercury
mmol: Millimole
mol: Mole
NCDs: Non-communicable diseases
PHCUs: Primary healthcare units
SBP: Systolic blood pressure
SNRS: Sidama National Regional State
T2DM: Type-2 diabetes mellitus
WHO: World Health Organization
WHO-PEN: World Health Organisation Package of Essential NCD
WTP: Willingness to pay

